# One-day preoperative systemic treatment regimen outcompetes five-day regimen in potentially resectable esophageal squamous cancer

**DOI:** 10.1101/2022.11.21.22282548

**Authors:** Sichao Wang, Shujie Huang, Zhen Gao, Yong Tang, Zihao Zhou, Guibin Qiao

**Affiliations:** Department of Thoracic Surgery, Guangdong Provincial People’s Hospital, Guangdong Academy of Medical Sciences, Guangzhou 510080, China

## Abstract

**Objective:** The current study aimed to compare the effectiveness of the one-day immunochemotherapy regimen, the one-day chemotherapy regimen and the five-day chemotherapy regimen in locally advanced esophageal squamous cell carcinoma patients treated with preoperative systemic treatment.

**Methods:** We retrospectively analyzed locally advanced ESCC patients who had received POST from January 2012 to September 2021 at the Department of Thoracic Surgery, Guangdong Provincial Peoples’ Hospital. The clinical and follow-up data were collected and analyzed according to 3 regimens. Categorical and continuous variables were analyzed by Chi-square and One-way ANOVA respectively, and overall survival (OS) was assessed using Kaplan-Meier analysis.

**Results:** A total of 395 POST-treated ESCC patients were enrolled, including 72 in the 5FU-based group, 168 in the pla/pac group, and 155 in the pla/pac/ICI group, and the mean follow-up time were 32.2, 44.2 and 14.3 months, respectively. the pla/pac/ICI group had the greatest benefit, with an ORR of 63.2% (P < 0.05) and a surgery conversion rate of 85.2% (P < 0.05). Furthermore, pla/pac/ICI group acquired a better short-term OS than the other groups (one-year OS: pla/pac/ICI 93.6% vs. pla/pac 87.4% vs. 5FU-based 70.5%).

**Conclusion:** In the context of the COVID-19 pandemic, one-day immunochemotherapy should be considered because it may yield higher response rates, bring better overall survival as well as greatly reduce the risk of treatment interruption. If immunotherapy is not available, the 1-day pla/pac regimen is also an effective and timely alternative.

## Introduction

The current standard-of-care treatment for locally advanced esophageal squamous cell carcinoma (ESCC) is preoperative chemoradiotherapy.^1^ However, most ESCC patients in China are from economically underdeveloped areas ^2^. The shortage of medical resources exhausts the willingness of such patients to receive radiotherapy. Therefore, neoadjuvant chemotherapy alone is a preferred option in Eastern Asia. The one-day platinum plus paclitaxel (pla/pac) regimen is commonly used in China, while the five-day platinum plus 5-fluorouracil (5FU-based) regimen is preferred in other countries.^3^ Especially in the context of the COVID-19 pandemic, how to deliver timely and effective treatment for cancer patients has become a huge challenge.^4^ Fortunately, The effectiveness of immunochemotherapy is encouraging. We compared the effectiveness of the one-day immunochemotherapy regimen, the one-day chemotherapy regimen and the five-day chemotherapy regimen in locally advanced ESCC patients treated with preoperative systemic treatment (POST).

## Methods

We retrospectively analyzed locally advanced ESCC patients who had received POST from January 2012 to September 2021 at the Department of Thoracic Surgery, Guangdong Provincial Peoples’ Hospital. The clinical and follow-up data were collected and analyzed according to 3 regimens including 5FU-based regimen which contains docetaxel+platinum+5-FU and platinum+5-FU regimens (5FU-based group), pla/pac regimen and pla/pac plus PD-1 inhibitor regimen (pla/pac/ICI). Complete response and partial response were defined as the objective response rate (ORR) per RECIST1.1 ^5^. Categorical and continuous variables were analyzed by Chi-square and One-way ANOVA respectively, and overall survival (OS) was assessed using Kaplan-Meier analysis. Statistical analysis was performed using SPSS v26 (Inc, Chicago, Illinois) and R, and a P value less than 0.05 was considered statistically significant.

## Results

A total of 395 POST-treated ESCC patients were enrolled, including 72 in the 5FU-based group, 168 in the pla/pac group, and 155 in the pla/pac/ICI group, and the mean follow-up time were 32.2, 44.2 and 14.3 months, respectively. There was no statistical difference in the baseline data among the three groups except POST cycles **(Table)**. As shown in the Figure 1A, the pla/pac/ICI group had the greatest benefit, with an ORR of 63.2% (P < 0.05) and a surgery conversion rate of 85.2% (P < 0.05). Moreover, the ypT0 or ypTis rate in the pla/pac/ICI group was significantly higher than that in 5FU-based and pla/pac groups. Furthermore, pla/pac/ICI group acquired a better short-term OS than the other groups (one-year OS: pla/pac/ICI 93.6% vs. pla/pac 87.4% vs. 5FU-based 70.5%, Figure 1B).

**Figure 1.**
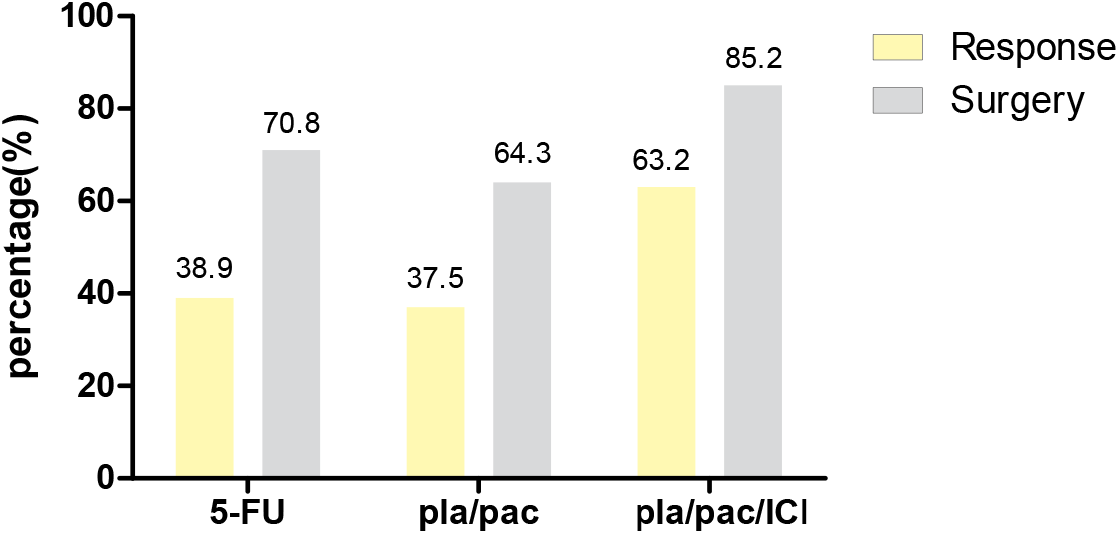

## Discussion

The COVID-19 pandemic has caused considerable consumption of medical resourse and cancer patients bear the brunt of delays in the interruption of medical care during the long treatment period^4^. Thus, it is urgent to deliver more effective and time-saving treatment. In this study, we demonstrated that the one-day pla/pac/IC group regimen has an obvious clinical advantage over one-day pla/pac and five-day 5FU-based regimen in terms of radiological/pathological tumor response, as well as long-term overall survival. Taken both time cost and efficacy into consideration, the one-day pla/pac/IC regimen migth be a more favorable option in treating locally advanced ESCC. Despite exciting results, patients treated with immunotherapy shoulder great financial burden. Our results showed that there were no significant differences in the surgical conversion rate and response rate between one-day pla/pac and five-day 5FU-based group. Furthermore, the pla/pac group showed better survival benefits than 5FU-based group. The one-day pla/pac regimen might be a better alternative until affordable immunotherapy could benefit the whole population.

## Conclusion

In the context of the COVID-19 pandemic, one-day immunochemotherapy should be considered because it may yield higher response rates, bring better overall survival as well as greatly reduce the risk of treatment interruption. If immunotherapy is not available, the 1-day pla/pac regimen is also an effective and timely alternative.

## Data Availability

All data produced in the present study are available upon reasonable request to the authors

## Acknowledgement

The study protocol was approved by the Ethics Committee of Guangdong Provincial People’s Hospital.

---

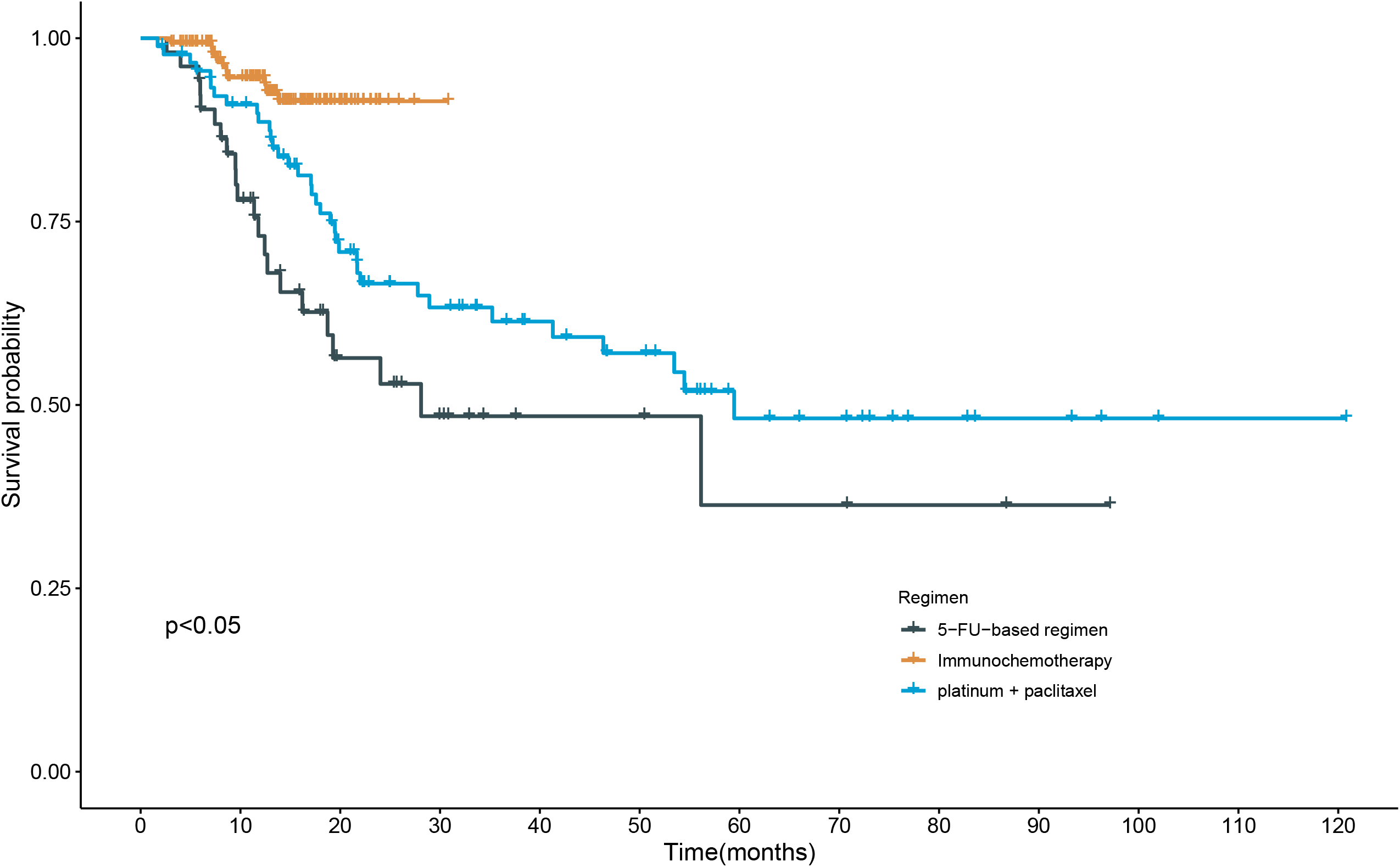

